# Trends in invasive and non-invasive *Streptococcus pneumoniae* disease in adults hospitalised in Bristol and Bath: a retrospective cohort study, 2006-2022

**DOI:** 10.64898/2025.12.12.25342129

**Authors:** Robert Challen, Catherine Hyams, David Hettle, Zahin Amin-Chowdhury, Charli Grimes, Gabriella Ruffino, Rauri Conway, Robyn Heath, Paul North, Adam Malin, Nick A Maskell, Philip Williams, O. Martin Williams, Shamez N Ladhani, Leon Danon, Adam Finn

## Abstract

**Objectives:** Pneumococcal surveillance typically focuses on invasive disease (IPD), collecting limited clinical data. We used a longitudinal adult cohort of IPD and non-invasive pneumococcal disease (non-IPD) patients in three hospitals between 2006-22, to describe clinical disease, including site of infection, and serotype distribution.

**Methods:** The demographics of IPD and non-IPD were compared and survival analysis performed. Features of IPD in different sites were compared and serotype distributions analysed using bootstrap resampling.

**Results:** Out of a total 3719 cases, non-IPD was responsible for 54.7%. IPD had a higher case-fatality ratio than non-IPD (14.8% versus 11.7%; P=0.005). Among the 1686 IPD cases, 15.3% were outside the respiratory tract. Those with meningitis were younger (54.9y vs LRTI 66.8y; P<0.001); and had high rates of ICU admission (63.4% vs LRTI 10.2%; P<0.001). We found a similar serotype distribution across different infection sites

**Conclusions:** Non-IPD accounts for 54.7% (2033/3719) of total diagnosed cases in adults hospitalised during the PCV era. IPD manifests outside of the respiratory tract in 15.3% (258/1686). Higher valent PCVs will likely have a similar impact regardless of different clinical presentations.

## BACKGROUND

*Streptococcus pneumoniae* remains a leading cause of mortality and morbidity worldwide, despite widespread use of effective pneumococcal vaccines^1^. The clinical disease spectrum in both children and adults is wide, ranging from asymptomatic carriage and mucosal infection, to bacteraemia and severe invasive disease including septicaemia and meningitis^2^. The clinical and economic burden of invasive pneumococcal disease (IPD) is particularly high in older adults, among whom IPD risk increases exponentially with patient age^1,3^. Additionally, case fatality rates remain high in this patient group; some evidence suggests that IPD disease in older adults may be becoming less severe in recent years^1^.

In England, the UK Health Security Agency (UKHSA) conducts national pneumococcal surveillance, reporting the serotype distribution of pneumococci isolated from sterile site culture^4,5^. Introduction of pneumococcal conjugate vaccines (PCVs) as part of the UK National Childhood Immunisation programme has resulted in diminution of circulation of PCV-serotypes through both direct and indirect effects, which has been followed by replacement with non-PCV serotypes in carriage and disease. In England, the introduction of PCV7 into the national childhood immunisation programme in 2006 resulted in near elimination of PCV7 serotypes in carriage and disease^6^. PCV7 replacement with PCV13 in 2010 led to further declines in IPD incidence, with PCV13-serotype IPD reducing from 4.46 cases per 100,000 population (combining all age groups) in 2008/09 to 1.66 cases per 100,000 population in 2016/17^5^. Over the same period, IPD due to non-PCV13 serotypes increased, especially in older adults, resulting in no net reduction in IPD cases in older adults in 2016/17 compared to pre-PCV13 incidence^5,7^. Importantly, UKHSA surveillance does not include non-invasive pneumococcal disease, such as that identified by urinary antigen testing (UAT). It is unclear whether some serotypes are more likely than others to cause disease at particular sites of infection and, since invasive potential varies between serotypes^8^, it is plausible that the distribution of causative pneumococcal serotypes may differ between invasive and non-invasive pneumococcal disease. Thus, current and future vaccines may have different coverage and effectiveness against IPD and non-invasive pneumococcal disease, and against infections at different body sites.

Most studies that describe the phenotype of pneumococcal disease in adults focus on invasive disease, particularly invasive lower respiratory tract infections, with few studies including non-invasive disease, and fewer still describing non-respiratory infections, such as meningitis, despite their global significance. Data on the clinical phenotype, disease severity, and causative serotypes in non-respiratory infections are especially sparse. In a national UK study, 20.7% laboratory-confirmed meningitis cases in adults aged 18-64 years and 27.9% of laboratory-confirmed meningitis cases in older adults were due to pneumococcus^9^, with some evidence of increasing incidence in recent years^10^. However, neither study included data on the responsible pneumococcal serotypes. In the Netherlands, a prospective cohort study comparing pre and post-PCV7 cohorts, found that although incidence and serotype distributions have changed, pneumococcal meningitis continues to have particularly poor outcomes^11^.

The objective of this study was to describe the epidemiology, clinical characteristics and outcomes of invasive and non-invasive pneumococcal disease over a 17-year period encompassing the PCV era in a retrospective cohort from a single geographical region in the UK. We also compared patient characteristics and outcomes of patients with invasive and non-invasive pneumococcal disease. Finally, focussing on the subgroup of patients with invasive disease, including meningitis, we compared patient characteristics, outcomes and serotype distributions across different infection sites.

Pneumococcal infection remains a significant global health concern, with significant associated morbidity and mortality, particularly among the very young, immunocompromised and older adults. While most studies provide disease incidence estimates, few are able to provide granular analyses concerning disease severity and clinical site of infection. The analyses undertaken here provide such insight, including the serotype distribution within meningitis, pneumonia and other sites of infection, and are thus important for informing policy decisions concerning vaccination and healthcare service provision globally.

## METHODS

### Study design

A retrospective, observational cohort study of all patients aged ≥16 years admitted to any of three large UK NHS hospitals (North Bristol, University Hospitals Bristol and Weston, and The Royal United Hospital NHS Trusts) in southwest England between 01/01/2006 and 31/12/2022 with microbiologically confirmed pneumococcal infection. These hospitals provide all secondary care within a defined geographical area, and include the regional cardiothoracic, pleural, respiratory specialist, and general medical and respiratory services and jointly have approximately 100,000 unplanned adult admissions annually.

Eligible cases were identified retrospectively by searching the Laboratory Information Management System (LIMS) database (Clinisys WinPath Enterprise). *S. pneumoniae* was identified by culture or PCR from a sterile site at a central laboratory using standard microbiological techniques combined with API®-20 Strep (BioMérieux, UK) or MALDI-TOF (matrix-assisted laser desorption/ionisation/time of flight) mass spectrometry (Bruker, UK). A positive pneumococcal urinary-antigen test [UAT] (BinaxNOW®, Alere, UK) was also considered confirmative of pneumococcal infection. Patients were included if they tested positive using any of these techniques. Confirmed cases were linked with the UKHSA national reference laboratory database to obtain serotype data, which were gathered at the end of the study to avoid risk of bias in clinical data collection. Clinical records were reviewed at each hospital and data were recorded in a standardised manner, including laboratory and radiological investigation results. Patient observations within 24 hours of clinical presentation with a pneumococcal infection were also recorded and vaccination status established using electronically-linked General Practitioner (GP) records. The CURB65 severity score^12^ on admission was calculated for each clinical episode. Clinical outcomes, including length of hospitalisation and Intensive Care Unit [ICU] admission, were also recorded. Inpatient mortality (i.e. patient death before discharge) was determined through review of the medical records. All adults were managed at the discretion of the admitting clinical team.

### Case definitions

Total pneumococcal disease included all positive cases whether identified by sterile site culture/PCR or positive UAT with clinical confirmation of infection. IPD was defined as either identification of pneumococcus from a sterile-site culture or the presence of a UAT positive test in a patient with a clinical diagnosis of infection in a sterile-site (e.g. meningitis, septic arthritis) with no other microbiological cause. Non-invasive pneumococcal disease was defined as positive UAT in patients without a clinical diagnosis of sterile site infection. The primary infection site was derived from the managing clinician’s diagnosis. Respiratory infection included both consolidative infection (i.e. pneumonia following BTS/NICE guidelines^13^) and non-pneumonic lower respiratory tract infection (LRTI). Pneumococcal serotypes were further grouped by vaccine-serotypes: PCV7, PCV13-7, PCV15-13, PCV20-15, PCV20-13, and serotypes not contained in a PCV (non-PCV) (Supplementary table 1). Patients’ pneumococcal risk was classified as “high risk” (i.e., immunocompromised), defined as those with chronic kidney disease, organ transplantation, immunodeficiency, haematological or solid malignancy, acquired immunodeficiency syndrome (AIDS), human immunodeficiency virus (HIV) infection, or treated with immunosuppressive drug therapy; “at-risk”, defined as those who did not have a high-risk condition, but who had asthma, congestive heart failure (CCF), chronic liver disease, COPD, diabetes mellitus, history of alcohol abuse, who were currently smokers or were over 65; or “low risk’’ in all other cases.

### Analysis

Data are reported as medians and interquartile ranges (IQR) for continuous variables, or means and standard deviations where their distribution was confirmed to be normal using the Anderson Darling normality test. Categorical variables are presented as counts and percentages. Multinomial confidence intervals were calculated using Wilson’s score interval method for binomial proportions^14^ which we tested on simulated data and found to be well calibrated for this problem. Baseline characteristics were compared using Fisher’s exact tests for categorical variables, and the two-sample Kolmogorov-Smirnov test for non-parametric continuous variables, Wilcoxon Rank Sum test for score variables, or 2 sided student’s t-test for parametric continuous variables^15^. We estimated survival probabilities using Cox proportional hazards regression and Kaplan-Meier curves to understand the impact of covariates on the survival outcomes^16^. We looked at end points of mortality, censored at the earliest of 30 days since admission or 31/12/2022, and discharge from hospital, censored at the earliest of 30 days since admission, patient death or 31/12/2022. For admission duration, increased likelihood of discharge implies shorter length of stay, and decreased likelihood of discharge implies longer length of stay. There were minimal missing data: for categorical variables, missing data were included as a separate category prior to statistical testing; for continuous variables, missing data were omitted for calculation of medians and interquartile ranges, and no statistical comparisons were made on items with missing values. There were no missing data in covariates in Cox regression models.

Pneumococcal disease incidence was estimated as the rate of monthly admission counts using a local polynomial regression, assuming a quasi-Poisson distribution, logistic link function, the R package ‘locfit’ according to the methods of Loader *et al.*^17^ using a bandwidth equivalent to 12 months’ worth of data and polynomial of degree one^18,19^.

Invasive pneumococcal disease consists of a number of clinical presentations including meningitis, which is known to have poor patient outcomes. The vast majority of non-invasive disease presents as lower respiratory tract disease. We conducted a sensitivity analysis, repeating our comparison of invasive versus non-invasive disease restricted to people with a presentation of lower respiratory tract disease alone.

In invasive disease, the serotype distributions of pneumococci responsible for different clinical presentations were compared by using a null hypothesis that serotype distribution is independent of clinical presentation. 10,000 bootstrap simulations of the same size as the sample were constructed. In each simulation, serotype and clinical presentation were randomly assigned using their observed marginal probability distributions and assuming independence. From the simulations the expected frequency of each combination of serotype and clinical presentation were estimated including empirical confidence intervals at 95% and 99.9%. Observed serotype counts for each clinical presentation were compared to expected values to identify combinations of serotype and clinical presentation observed unexpectedly frequently or infrequently.

All analyses were performed using R version 4.2^20^, and data flow diagrams generated using the R package ‘dtrackr’^21^.

## RESULTS

During 2006-2022, we identified 3719 adults with pneumococcal disease, including 1686 with IPD and 2033 with non-invasive pneumococcal disease. Amongst IPD cases, 84.7% (1428/1686) were respiratory infection (LRTI, Effusion or Empyema), 10.2% (172/1686) had meningitis, 2.0% (34/1686) had septic arthritis and 3.1% (52/1686) were infections at other sites (Figure 1). There were no statistically significant differences in age, sex, smoking status or recreational drug use between patients hospitalised with IPD and non-invasive pneumococcal disease, but chronic underlying respiratory and cardiac conditions were more common in those hospitalised with IPD (Table 1).

**Figure 1:**
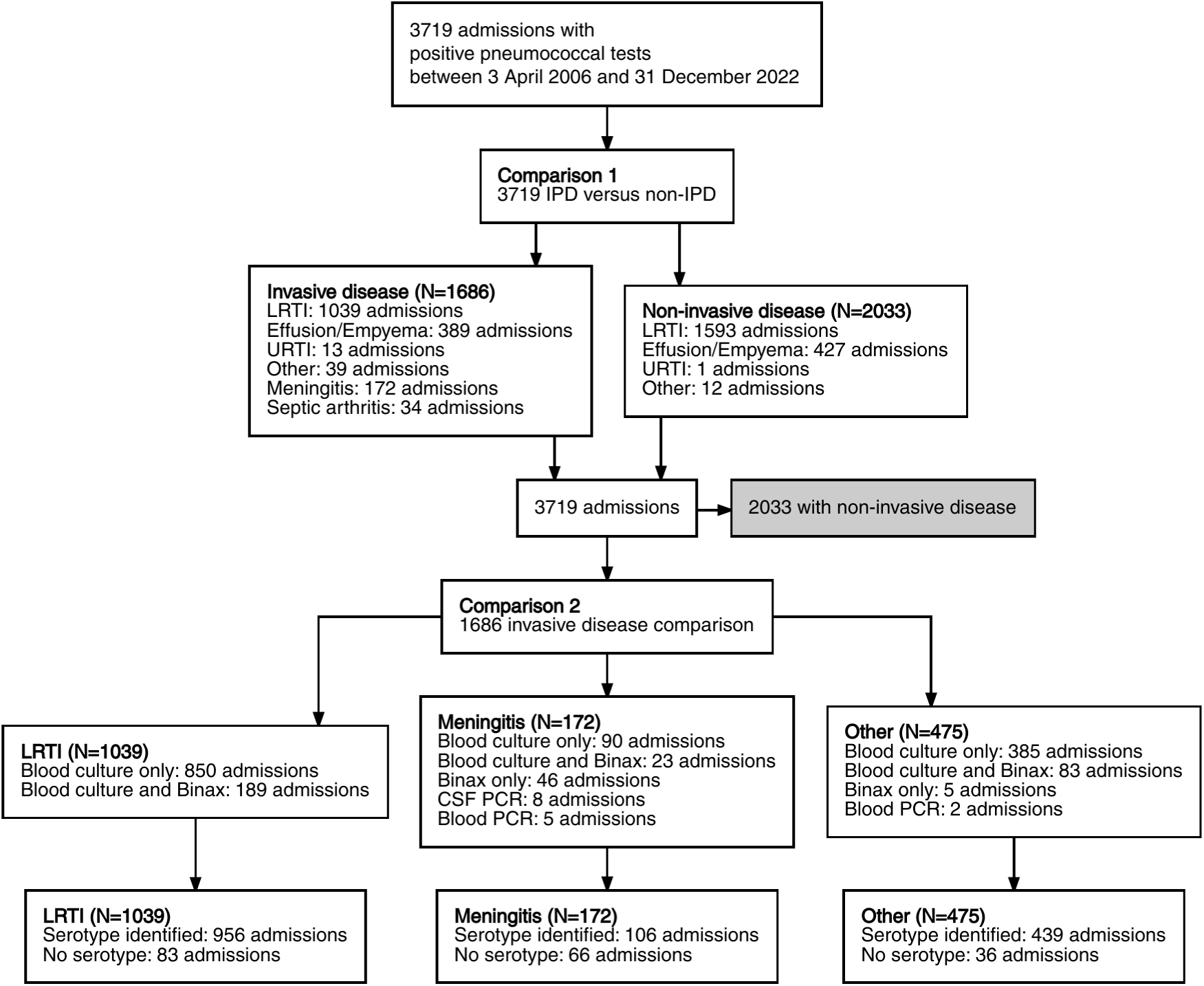
Data flow diagram. The main analysis focuses on two comparisons: (1) between invasive and non-invasive pneumococcal disease, and (2) within the IPD disease cohort we compare clinical presentations, classified as lower respiratory tract infection (LRTI, both pneumonic and non-pneumonic lower respiratory tract infection); meningitis; and other causes, including septic arthritis and other sites of infection.

**Table 1:** Demographic characteristics of patients with IPD versus non-invasive pneumococcal disease.

| Variable | Characteristic | Non-invasive disease<br>(N=2033) | IPD<br>(N=1686) | P value |
| --- | --- | --- | --- | --- |
| Age | Median [IQR] | 66.3 [50—78.8] | 65.9 [50.8—79] | 0.74 † |
| Gender | Female % (n) | 51.2% (1040) | 49.8% (839) | 0.41 †† |
| Pneumococcal Risk Classification | Low risk % (n) | 10.2% (207) | 8.8% (148) | 0.14 †† |
|  | At risk % (n) | 65.6% (1334) | 64.7% (1091) |  |
|  | High risk % (n) | 24.2% (492) | 26.5% (447) |  |
| Smoker | Non-smoker % (n) | 29.4% (597) | 29.4% (495) | 0.95 †† |
|  | Ex-smoker % (n) | 40.8% (829) | 40.3% (680) |  |
|  | Current % (n) | 29.9% (607) | 30.3% (511) |  |
| Alcohol Abuse | yes % (n) | 7.9% (160) | 7.3% (123) | 0.53 †† |
| CCI Category | None (0) % (n) | 17.3% (352) | 17.1% (289) | 0.15 †† |
|  | Mild (1-2) % (n) | 18.8% (382) | 19.4% (327) |  |
|  | Moderate (3-4) % (n) | 26.1% (531) | 23.1% (389) |  |
|  | Severe (5+) % (n) | 37.8% (768) | 40.4% (681) |  |
| Any Chronic Lung Disease | yes % (n) | 42.4% (862) | 35.8% (604) | <0.001 †† |
| Any Chronic Heart Disease | yes % (n) | 46.7% (950) | 50.2% (847) | 0.035 †† |
| COPD | yes % (n) | 29.6% (602) | 25.4% (429) | 0.0052 †† |
| PPV23 Vaccination | no % (n) | 57.6% (1171) | 57.4% (967) | 0.93 †† |
|  | yes % (n) | 42.4% (861) | 42.6% (719) |  |
|  | <missing> % (n) | 0.0% (1) | 0.0% (0) |  |
| White Cell Count | Median [IQR] | 16.1<br>[11.3—21.8] | 16.4 [11.5—22.2] | — ††† |
| CRP | Median [IQR] | 189<br>[106—309] | 196<br>[115—312] | — ††† |

†, 2 sample Kolmogorov-Smirnov test (continuous); ††, Fisher's exact test (categorical); †††, Not calculated due to missing values (continuous)
Normal distributions determined by the Anderson-Darling test ( $P > 0.005$ ) An adjusted P value of 0.00417 may be considered significant.
CCI, Charlson Comorbidity Index; CRP, C-reactive protein; COPD, chronic obstructive pulmonary disease; IPD, invasive pneumococcal disease; IQR, interquartile range; n, number; PPV23, 23-valent pneumococcal polysaccharide vaccine

IPD was associated with worse clinical outcomes than non-invasive pneumococcal disease (30-day case fatality rates (CFR), 14.8% (250/1686) versus 11.7% (238/2033); P 0.0054 and greater requirement for ventilation, 14.0% (236/1686) versus 11.5% (234/2033); P 0.026, (Table 2). When adjusted for age, sex and chronic lung disease, IPD was associated with a significantly increased hazard ratio (HR [95% confidence interval - CI] 1.29 [1.08 – 1.55]; P 0.005) for 30-day CFR compared to non-invasive pneumococcal disease (Table 3 and Figure 2; sensitivity analysis in Supplementary table 3). IPD was also associated with a reduced likelihood of hospital discharge, (HR 0.92 [0.86 – 0.99]; P 0.022), equating to a longer inpatient stay, compared to non-invasive pneumococcal disease in the main analysis (Table 3) but this was not significant in the subset of the cohort with lower respiratory tract disease only (Supplementary table 4).

**Table 2:** Clinical outcomes of patients with IPD versus non-invasive pneumococcal disease.

| Variable | Characteristic | Non-invasive disease<br>(N=2033) | IPD<br>(N=1686) | P value |
| --- | --- | --- | --- | --- |
| CURB65 Category | 0-1 (Mild) % (n) | 36.9% (751) | 34.2% (576) | 0.002 † |
|  | 2 (Moderate) % (n) | 31.9% (649) | 29.0% (489) |  |
|  | 3-5 (Severe) % (n) | 31.1% (633) | 36.8% (621) |  |
| Admitted ICU | yes % (n) | 16.4% (333) | 17.9% (301) | 0.24 †† |
| Intubation | yes % (n) | 11.5% (234) | 14.0% (236) | 0.026 †† |
| Received Inotropes | yes % (n) | 13.0% (264) | 13.9% (235) | 0.41 †† |
| Death Within 30 Days | yes % (n) | 11.7% (238) | 14.8% (250) | 0.0054 †† |

†, Chi-squared Test for Trend in Proportions (ordered); ††, Fisher's exact test (categorical)
An adjusted P value of 0.01 may be considered significant.
ICU, intensive care unit; IPD, invasive pneumococcal disease.

**Table 3:** Adjusted Cox proportional hazard models for patient outcomes. The outcomes of death and discharge from hospital within 30 days (censored for death) are adjusted for by factors that differ significantly between groups including age, gender, and chronic lung disease. In the analysis for death, an increased hazard implies a poorer outcome; whereas, in the analysis for hospital discharge, an increased probability of discharge is equivalent to a reduced length of stay, and lower hazard rate implies a poorer outcome.

| Characteristic | Subgroup | Death within 30 days (N=3060) |  | Discharge within 30 days (N=3060) |  |
| --- | --- | --- | --- | --- | --- |
|  |  | HR [95% CI] | P value | HR [95% CI] | P value |
| Invasive Status | Non-invasive disease | ref | 0.005 | ref | 0.022 |
|  | Invasive disease | 1.29 [1.08 – 1.55] |  | 0.92 [0.86 – 0.99] |  |
| Age Category | 18-34 | ref | <0.001 | ref | <0.001 |
|  | 35-49 | 1.40 [0.71 – 2.75] |  | 0.89 [0.78 – 1.03] |  |
|  | 50-64 | 2.43 [1.32 – 4.48] |  | 0.72 [0.63 – 0.82] |  |
|  | 65-74 | 4.14 [2.27 – 7.54] |  | 0.69 [0.60 – 0.78] |  |
|  | 75-84 | 5.37 [2.97 – 9.74] |  | 0.58 [0.51 – 0.67] |  |
|  | 85+ | 10.19 [5.65 – 18.36] |  | 0.48 [0.41 – 0.56] |  |
| Gender | Female | ref | 0.003 | ref | 0.005 |
|  | Male | 1.32 [1.10 – 1.58] |  | 0.90 [0.84 – 0.97] |  |
| Any Chronic Lung Disease | no | ref | 0.137 | ref | 0.172 |
|  | yes | 1.15 [0.96 – 1.38] |  | 1.05 [0.98 – 1.13] |  |

P values calculated using Likelihood ratio test (II)

**Figure 2:**
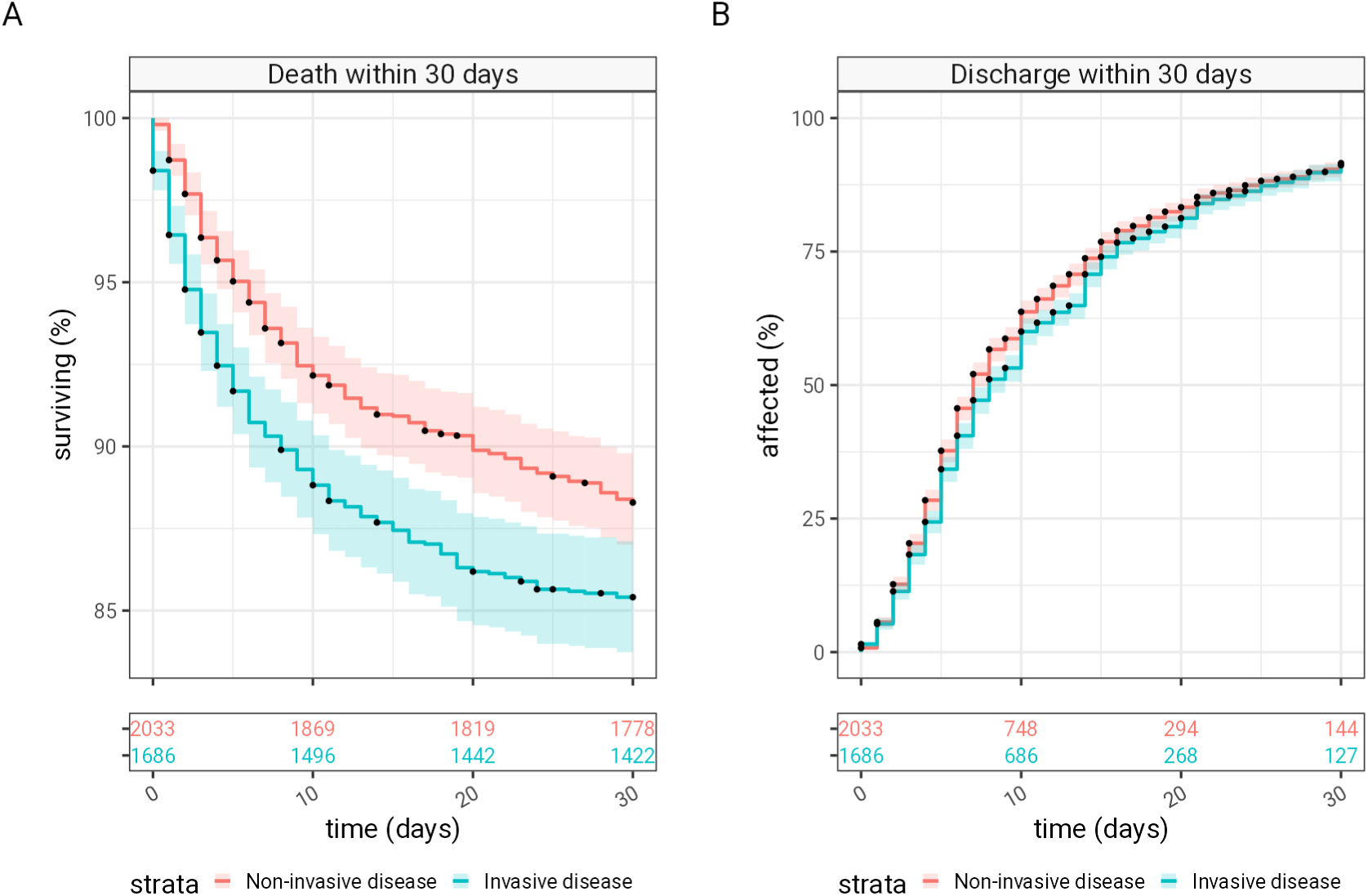
Kaplan-Meier survival analysis for patients with pneumococcal disease. Survival analysis for patients hospitalised with invasive (blue) versus non-invasive (red) pneumococcal disease for (A) death and (B) discharge within the first 30 days (with discharge censored by inpatient death). In both graphs, the number of patients at risk is shown in the table underneath each plot. 95% confidence intervals are represented with shaded bars. Note that the scale of the y-axis on panel A does not include zero.

Among 1686 patients with IPD, we identified 172 patients with meningitis, 1039 with lower respiratory tract infection and 475 with infection at another clinical site (Figure 1, Table 4). Compared to those with respiratory and other site infection, patients with meningitis were younger, had fewer pre-existing medical comorbidities and were less likely to smoke (all P<0.001) (Table 4). Pneumococcal meningitis was associated with the same inpatient survival as those with other types of IPD (P=0.56) but with higher rates of ICU admission, intubation and inotropic support (all P<0.001) (Table 5). Supplementary table 5 compares factors that are associated with survival in those with meningitis versus those that affect survival in other IPD groups. While age is the dominant factor associated with survival in LRTI, it was not significantly correlated to outcome in meningitis, which had statistical associations with comorbidities (including COPD) and male gender.

**Table 4:** Demographics of patients with IPD, classified by site of infection. Demographic characteristics of patients with IPD comparing the pneumococcal disease presentation, classified as lower respiratory tract infection (LRTI, both pneumonic and non-pneumonic lower respiratory tract infection); meningitis; and, other causes, including septic arthritis, and other sites of infection.

| Variable | Characteristic | LRTI<br>(N=1039) | Meningitis<br>(N=172) | Other<br>(N=475) | P value |
| --- | --- | --- | --- | --- | --- |
| Age | Median [IQR] | 66.8 [52—79.6] | 54.9 [39.2—65.6] | 68.7 [53.2—80.7] | <0.001 † |
| Gender | Female % (n) | 49.5% (514) | 48.8% (84) | 50.7% (241) | 0.87 †† |
| Pneumococcal Risk Classification | Low risk % (n) | 6.8% (71) | 28.5% (49) | 5.9% (28) | <0.001 †† |
|  | At risk % (n) | 65.9% (685) | 56.4% (97) | 65.1% (309) |  |
|  | High risk % (n) | 27.2% (283) | 15.1% (26) | 29.1% (138) |  |
| Smoker | Non-smoker % (n) | 27.5% (286) | 49.4% (85) | 26.1% (124) | <0.001 †† |
|  | Ex-smoker % (n) | 43.7% (454) | 21.5% (37) | 39.8% (189) |  |
|  | Current % (n) | 28.8% (299) | 29.1% (50) | 34.1% (162) |  |
| Alcohol Abuse | yes % (n) | 5.9% (61) | 5.8% (10) | 10.9% (52) | 0.0021 †† |
| CCI Category | None (0) % (n) | 16.2% (168) | 32.6% (56) | 13.7% (65) | <0.001 †† |
|  | Mild (1-2) % (n) | 17.1% (178) | 32.6% (56) | 19.6% (93) |  |
|  | Moderate (3-4) % (n) | 24.5% (255) | 18.6% (32) | 21.5% (102) |  |
|  | Severe (5+) % (n) | 42.2% (438) | 16.3% (28) | 45.3% (215) |  |
| Any Chronic Lung Disease | yes % (n) | 39.7% (412) | 19.8% (34) | 33.3% (158) | <0.001 †† |
| Any Chronic Heart Disease | yes % (n) | 51.7% (537) | 30.8% (53) | 54.1% (257) | <0.001 †† |
| COPD | yes % (n) | 28.3% (294) | 9.9% (17) | 24.8% (118) | <0.001 †† |
| PPV23 Vaccination | yes % (n) | 43.9% (456) | 21.5% (37) | 47.6% (226) | <0.001 †† |
| White Cell Count | Median [IQR] | 16.2 [11.3—21.5] | 17.3 [13.4—24.5] | 16.4 [11.4—22.7] | — ††† |
| CRP | Median [IQR] | 189 [114—310] | 192 [98—306] | 206 [128—341] | — ††† |

†, Kruskal-Wallis rank sum test (continuous); ††, Fisher's exact test (categorical); †††, Not calculated due to missing values (continuous)
Normal distributions determined by the Anderson-Darling test ( $P>0.005$ ) An adjusted P value of 0.00417 may be considered significant.
CCI, Charlson Comorbidity Index; CRP, C-reactive protein; COPD, chronic obstructive pulmonary disease; LRTI, lower respiratory tract infection; IPD, invasive pneumococcal disease; IQR, interquartile range; n, number; PPV23, 23-valent pneumococcal polysaccharide vaccine

**Table 5:** Clinical outcomes of the patients with IPD, stratified by site of infection.

| Variable | Characteristic | LRTI<br>(N=1039) | Meningitis<br>(N=172) | Other<br>(N=475) | P value |
| --- | --- | --- | --- | --- | --- |
| CURB65 Category | 0-1 (Mild) % (n) | 33.4% (347) | 43.0% (74) | 32.6% (155) | 0.012 † |
|  | 2 (Moderate) % (n) | 28.8% (299) | 32.0% (55) | 28.4% (135) |  |
|  | 3-5 (Severe) % (n) | 37.8% (393) | 25.0% (43) | 38.9% (185) |  |
| Admitted ICU | yes % (n) | 10.2% (106) | 63.4% (109) | 18.1% (86) | <0.001 † |
| Intubation | yes % (n) | 7.2% (75) | 52.3% (90) | 14.9% (71) | <0.001 † |
| Received<br>Inotropes | yes % (n) | 7.9% (82) | 48.8% (84) | 14.5% (69) | <0.001 † |
| Death Within 30<br>Days | yes % (n) | 15.4% (160) | 12.2% (21) | 14.5% (69) | 0.56 † |

†, Fisher's exact test
An adjusted P value of 0.01 may be considered significant.
ICU, intensive care unit, LRTI, lower respiratory tract infection

There were no significant differences in the distribution of pneumococcal serotypes responsible for respiratory infection, meningitis and other IPD (Figure 3), after adjusting for multiple testing (Supplementary figure 2), with no evidence of statistically significant differences in the serotype distributions at P<0.001 (Supplementary figure 3). The temporal pattern in meningitis cases also followed that for respiratory infection and other IPD. IPD cases have been on an upward trend between 2009 and 2019 (figure 4). Overall IPD cases fell during the first year of the COVID-19 pandemic and then started to return to pre-pandemic levels; the trajectory of the tail of the time series must be interpreted with caution due to limited information constraining the model at this point (Figure 4).

**Figure 3:**
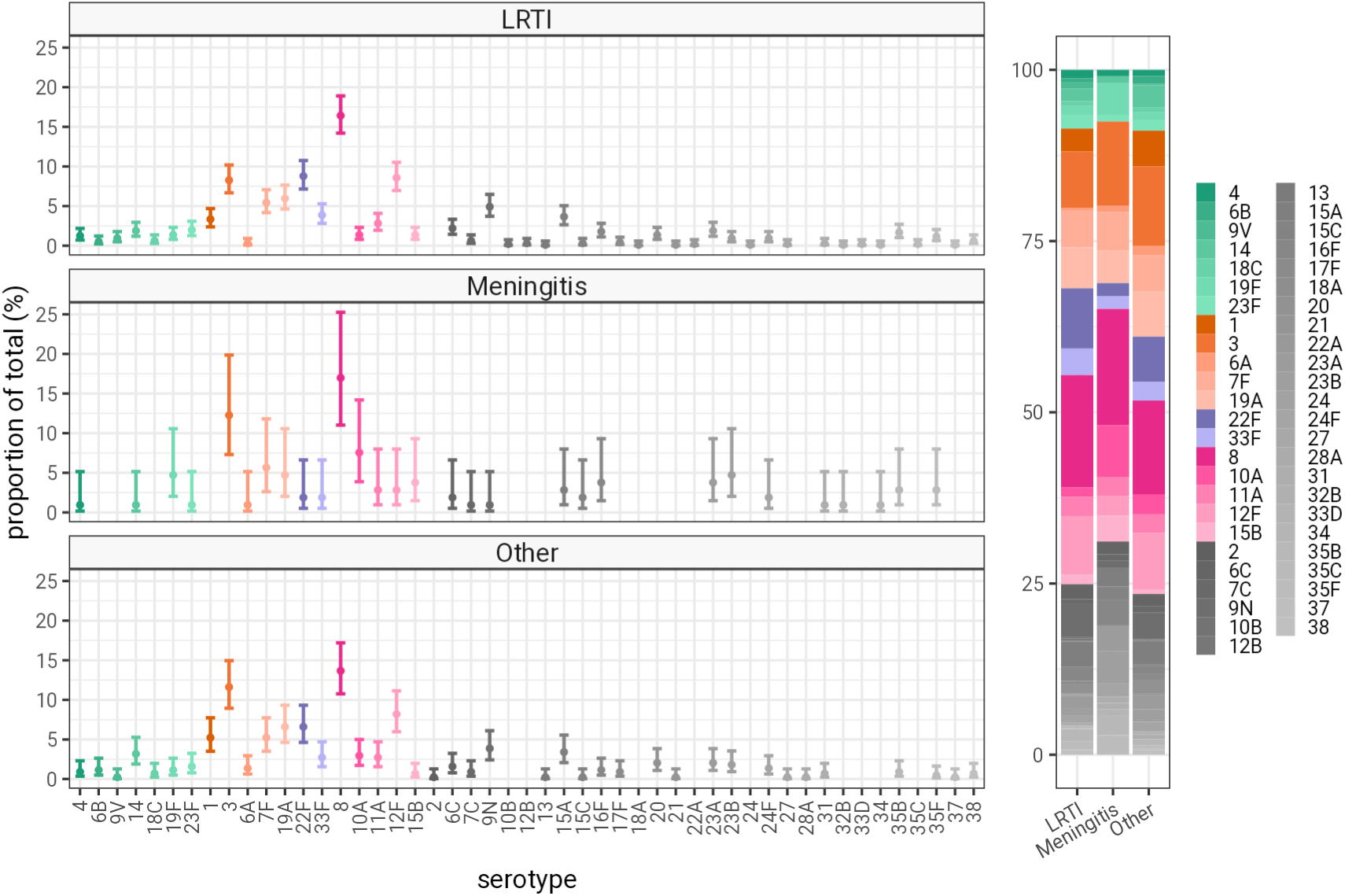
Pneumococcal serotype distribution of IPD, stratified by site of infection. The serotype distribution for pneumococcal cases where a serotype was identified,stratified by pneumococcal site of infection (classified as LRTI [lower respiratory tract infection, including both pneumonic and non-pneumonic respiratory infection]; meningitis; and, other causes such as septic arthritis, etc). Major colours represent polysaccharide capsular vaccine types, PCV7 in green, PCV13 in orange, PCV15 in violet, and PCV20 in magenta. Non-PCV vaccine serotypes are in grey. Serotypes which were not observed are not displayed (including PCV13 serotype 5). Proportion estimates are plotted with binomial 95% confidence intervals.

**Figure 4:**
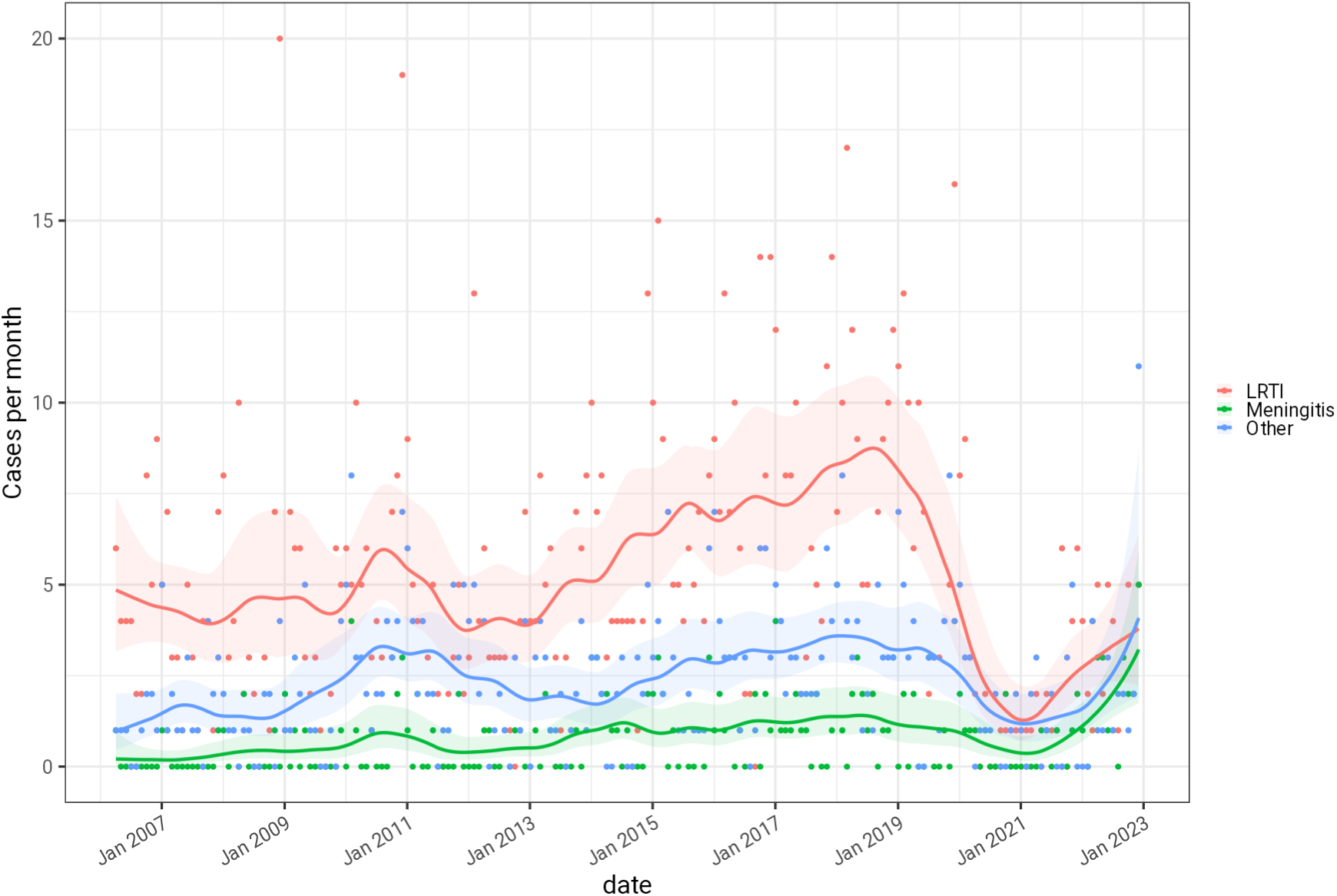
The temporal distribution of IPD cases in Bristol, 2006-2022. Cases are stratified by pneumococcal disease presentation: LRTI, meningitis, and other causes - including effusions, empyema, and septic arthritis. Solid lines show a modelled case rate per month based on a poisson process, with 95% confidence intervals. The trajectory of the tail of the time series should not be over-interpreted as it is operating on limited information.

## DISCUSSION

This analysis of pneumococcal disease over 17 years provides evidence that IPD remains associated with worse clinical outcomes than non-invasive pneumococcal disease in adults. Importantly, this study compares the severity of invasive and non-invasive disease, and severity of disease by clinical site of infection, as well as report the serotypes responsible for invasive disease. IPD was associated with a 29% increase in the risk of death within 30 days after controlling for patient age and gender, and chronic lung disease. Our data show that 54.7% (2033/3719) of pneumococcal disease in hospitalised adults is non-invasive, and the remainder invasive. This disease burden is not accounted for in studies reporting only IPD cases and may not be recognised in studies focused on pneumococcal cases identified through sterile site culture or PCR alone. While IPD was more severe than non-invasive pneumococcal disease, there remains substantial morbidity and mortality in patients hospitalised with non-invasive pneumococcal disease, with 11.7% (238/2033) of patients dying within 30 days of diagnosis. Hospital length of stay is only marginally longer (reported above as reduced risk of discharge) in patients with IPD compared to non-invasive pneumococcal disease, and this difference disappeared when meningitis cases were excluded from the analysis.

During 2009-2019, there was an upward trend in pneumococcal meningitis cases (Figure 4). This finding concurs with a literature review conducted in 2019, that concluded that there had been an initial decline in pneumococcal vaccine-serotype meningitis following PCV introduction in Europe and North America, but this temporary decline was reversed in several regions due to serotype replacement by non-vaccine serotypes^22^. While some countries including Finland, the USA, Israel and Germany have seen declines in pneumococcal meningitis after PCV7 and PCV13 implementation^23,24,25,26^, there were reports of an increase in meningitis caused by non-PCV serotypes^27^ including a French study that reported a dip in incidence following PCV7 implementation followed by rebound in pneumococcal meningitis in infants caused by non-PCV serotypes^28^. The overall increase in meningitis incidence is also a feature of recent UK national data on laboratory confirmed meningitis cases^10^, in which annual pneumococcal meningitis incidence increased by 0.02/100,000 per year (95% CI, 0.01–0.04/100,000) from 2012-19 in the context of significant declines in meningitis due to Group A Streptococcus, meningococcus and *Mycobacterium tuberculosi*s. During the COVID-19 pandemic (Dec 2019 through to Dec 2021), the incidence of pneumococcal meningitis fell, presumably due to social distancing measures, but this was soon followed by a resurgence during 2022 and 2023 when pandemic restrictions were removed, with incidence exceeding pre-pandemic levels. Further data are needed to confirm whether this is a transient post-pandemic phenomenon, but this trend is of concern given the severe outcomes of pneumococcal meningitis in young patients reported in this and other cohorts^9,11^.

The demographics, patient characteristics and outcomes for pneumococcal meningitis were different from those for pulmonary infection. Meningitis patients were younger and less frequently vaccinated with PPV23 than those with respiratory infection. Our results are in keeping with a national cohort study conducted in the Netherlands, which found that unfavourable outcome and case fatality rates increased with age, but this study did not account for vaccination status of the patients^11^. In the UK, PPV23 vaccination is offered to adults aged ≥65y or those ≥2 years with underlying conditions that increase their risk of pneumococcal disease^29^. As such, these data do not allow us to determine whether age or vaccination are independently correlated with pneumococcal meningitis. In keeping with national surveillance, we report a 30 day CFR of 12.2% (21/172) in pneumococcal meningitis, which is similar to the 16.5% reported for laboratory-confirmed pneumococcal meningitis nationally^10^, both of which are lower than the 20% in-hospital CFR reported in the Netherlands by Koelman *et al*^11^. Given the high morbidity, mortality and healthcare use, including ICU admission, for pneumococcal meningitis, increasing public awareness, improving clinical management, developing new therapeutic options for pneumococcal meningitis, and monitoring the effect of pneumococcal vaccination programmes are all indicated.

This analysis contributes to the global literature by describing the serotype distribution of pneumococci responsible for respiratory infection, meningitis, and other clinical infections within the same cohort of adult patients in the era of routine PCV use in children. This analysis has global public health importance, and importantly we did not identify any differences between the serotype distributions of pneumococci causing IPD at different clinical infection sites. Broadly, the serotypes causing respiratory infection aligned with those causing meningitis and infections at other sites. However, minor differences in serotype distribution may not have been picked up in our study because of our relatively small sample size for some of the sites of pneumococcal infections. Nevertheless, the similar distributions of pneumococcal serotypes implies that new vaccines should have a similar impact in reducing pneumococcal disease burden due to vaccine serotypes affecting all clinical presentations, including meningitis. Laboratory-based surveillance in the UK and other countries for pneumococcal meningitis does not contain any serotype information, but the national increase in laboratory-confirmed pneumococcal meningitis cases aligns with increasing IPD cases in the UK, which has disproportionately affected adults more than children^5^, probably because of serotype replacement, initially in carriage in young children and then in disease in adults and older adults. National surveillance reports fluctuating serotype 3 incidence during 2000-17, but by 2018 this serotype was the third most common serotype responsible for pneumococcal disease, after serotype 8 and 12F^5^, with serotypes 3 and 8 being responsible for 13% and 17% of IPD, respectively, in adults aged ≥65 years^7^. In concordance with previous results from this cohort^19^, we found the majority of IPD cases that presented as lower respiratory tract infection, meningitis and other site infections were attributable to serotype 3 or 8. Previous reports have suggested that serotype 3 and 8 may have a propensity to cause meningitis ^9,10^; in contrast, we found no difference between the proportions of pneumococcal respiratory infection and of meningitis attributable to these serotypes in our cohort.

This analysis benefits from including a large retrospective cohort of invasive and non-invasive pneumococcal disease with detailed clinical information, including both risk factors and outcomes. In addition, linkage with the UKHSA reference laboratory enabled us to report serotype data where available. As this is a hospitalised cohort there is a potential bias resulting from selection on admission or case identification, and it is possible that a lower threshold for investigation of some patient groups, such as those with comorbidities or frailty, would result in increased detection of relatively less severe cases in these at-risk groups; fewer similar cases would be detected in groups not at-risk and hence not prioritised for investigation. This would tend to reduce the observed impact of frailty or age on severity. Identification of non-invasive disease in this cohort has increased over time^19^ as BinaxNOW urine antigen testing has become more widely used. There are likely to be fewer non-invasive cases identified before 2009, and the non-invasive cases identified may be disproportionately more severe at that time. This could make the non-invasive cases appear more severe, but this effect would only be seen for a small part of the data set. This regional study might not be representative of other regions or populations, although reassuringly, the data are concordant with national UKHSA epidemiological and BTS pneumonia audit data^30^. The study also does not include pneumococcal infections in the community which may be successfully treated in primary care without the need for hospital investigation, management or treatment.

In this study, we found that non-invasive pneumococcal disease was responsible for 54.7% (2033/3719) of total pneumococcal disease in a hospitalised cohort of patients over a period of 17 years in Southwest England. Case fatality rates were higher for IPD compared to non-invasive pneumococcal disease, although the latter was also associated with a 30-day CFR of 11.7% (238/2033). Therefore, the true burden of pneumococcal disease, including its impact on healthcare resources, may be significantly underestimated when only invasive disease is accounted for, which will have implications for public health planning and provision of healthcare resources globally. Adults with pneumococcal meningitis were typically younger than other pneumococcal infections and had a very high risk of intensive care admission, but similar in-hospital fatality rates to other pneumococcal presentations. Pneumococcal serotypes causing invasive disease did not differ by clinical presentation, with serotypes 3 and 8 dominating IPD across all subgroups of disease. Higher valency PCVs will probably have a similar impact regardless of different clinical presentations and given their recent licensure, the impact of future vaccination policy changes in the UK and other countries, including changes to the paediatric immunisation programmes and increased use of conjugate vaccines in adults, will also need careful evaluation.

## DECLARATIONS

### Ethics approval and consent to participate

This study was approved by the Health Research Authority, UK (IRAS 265437).

### Availability of data and materials

The data used in this study are sensitive and cannot be made publicly available without breaching patient confidentiality rules. Therefore, individual participant data and a data dictionary is not available to other researchers.

### Authors’ contributions

CH, RCh, OMW, SL and AF generated the research questions and analysis plan. CH, RH, DH, GF, RCo, PN, PW, ZAC collected data. CH, RH, and DH verified the data. CH, RCh, LD, and AF undertook the data analysis. AF, OMW and AM provided oversight of the research. All authors contributed to the preparation of the manuscript and its revision for publication and had responsibility for the decision to publish.

## Acknowledgements

The authors would like to acknowledge the research teams at The Royal United, North Bristol and University Hospitals of Bristol and Weston NHS Trusts.

