## Supplementary material for "Trends in invasive and non-invasive *Streptococcus pneumoniae* disease in adults hospitalised in Bristol and Bath: a retrospective cohort study, 2006-2022"

**Contents:**

| | |
|---|---|
| Vaccine-type serotype groupings | 2 |
| Sensitivity analysis: Invasive versus non-invasive disease in the LRTI cohort. | 3 |
| Survival analysis: Invasive pneumococcal disease stratified by clinical presentation | 6 |
| Serotype distributions in invasive pneumococcal disease | 9 |

#### Vaccine-type serotype groupings

*Supplementary table 1: Serotypes contained within each pneumococcal vaccination and vaccine group.*

| Vaccination | Serotypes |
|---|---|
| PPV-23, PneumoVax® | 1, 2, 3, 4, 5, 6B, 7F, 8, 9N, 9V, 10A, 11A, 12F, 14, 15B, 17F, 18C, 19F, 19A, 20, 22F, 23F, 33F. |
| PCV-7, Prevenar® | 4, 6B, 9V, 14, 18C, 19F and 23F |
| PCV-13, Prevenar13® | 1, 3, 4, 5, 6A, 6B, 7F, 9V, 14, 18C, 19A, 19F, & 23F |
| PCV-15, VAXNEUVANCE® | 1, 3, 4, 5, 6A, 6B, 7F, 9V, 14, 18C, 19A, 19F, 22F, 23F and 33F |
| PCV-20, Prevenar20® | 1, 3, 4, 5, 6A, 6B, 7F, 8, 9V,10A, 11A, 12F, 14, 15B, 18C, 19A, 19F, 22F, 23F and 33F |
| PCV13-7 serotypes | 1, 3, 5, 6A, 7F, 19A |
| PCV15-13 serotypes | 22F, 33F |
| PCV20-15 serotypes | 8, 10A, 11A, 12F, 15B |
| PCV20-13 serotypes | 8, 10A, 11A, 12F, 15B, 22F, 33F |
| Non-PCV serotypes | Any serotype not contained in PCV20 (and therefore PCV15, PCV13 and PCV7) |

PCV, Pneumococcal conjugate vaccine; PPV-23, Pneumococcal polysaccharide vaccine, 23 valent

### Sensitivity analysis: Invasive versus non-invasive disease in the LRTI cohort.

As the effect of meningitis is significantly different to that of LRTI and by its nature restricted to the invasive disease cohort this could influence our findings. A sensitivity analysis comparing invasive versus non-invasive disease in the LRTI presentation category only removes this potential bias.

*Supplementary table 2: Demographic characteristics of invasive versus non invasive disease in the subgroup of patients with LRTI.*

| Variable | Characteristic | Non-invasive LRTI Value (N=1593) | Invasive LRTI Value (N=1039) | P value |
|---|---|---|---|---|
| Age | Median [IQR] | 66 [49.7—78.5] | 66.8 [52—79.6] | 0.24 † |
| Gender | Female % (n) | 52.9% (842) | 49.5% (514) | 0.094 †† |
| Pneumococcal Risk Classification | Low risk % (n) | 11.0% (176) | 6.8% (71) | <0.001 †† |
| | At risk % (n) | 66.1% (1053) | 65.9% (685) | |
| | High risk % (n) | 22.8% (364) | 27.2% (283) | |
| Smoker | Non-smoker % (n) | 30.9% (493) | 27.5% (286) | 0.1 †† |
| | Ex-smoker % (n) | 40.0% (637) | 43.7% (454) | |
| | Current % (n) | 29.1% (463) | 28.8% (299) | |
| Alcohol Abuse | yes % (n) | 8.1% (129) | 5.9% (61) | 0.031 †† |
| CCI Category | None (0) % (n) | 17.8% (283) | 16.2% (168) | 0.063 †† |
| | Mild (1-2) % (n) | 19.1% (304) | 17.1% (178) | |
| | Moderate (3-4) % (n) | 26.2% (418) | 24.5% (255) | |
| | Severe (5+) % (n) | 36.9% (588) | 42.2% (438) | |
| Any Chronic Lung Disease | yes % (n) | 43.4% (691) | 39.7% (412) | 0.063 †† |
| Any Chronic Heart Disease | yes % (n) | 45.6% (726) | 51.7% (537) | 0.0024 †† |
| COPD | yes % (n) | 29.9% (477) | 28.3% (294) | 0.38 †† |
| PPV23 Vaccination | yes % (n) | 42.1% (671) | 43.9% (456) | 0.38 †† |
| White Cell Count | Median [IQR] | 15.9 [11.3—21.4] | 16.2 [11.3—21.5] | — ††† |
| CRP | Median [IQR] | 186 [104—300] | 189 [114—310] | — ††† |

†, 2 sample Kolmogorov-Smirnov test (continuous); ††, Fisher's exact test (categorical); †††, Not calculated due to missing values (continuous)
Normal distributions determined by the Anderson-Darling test (P>0.005)
An adjusted P value of 0.00417 may be considered significant.

There are few large clear differences between invasive and non-invasive disease cohorts in the subgroup with LRTI. People with invasive disease tend to be in the high pneumococcal risk category, and have more chronic heart disease. These trends are broadly in line with the comparison of invasive to non-invasive disease of all clinical presentations in table 1 of the main paper.

*Supplementary Table 3: Comparison of outcomes of invasive versus non invasive disease in the subgroup of patients with LRTI.*

| Variable | Characteristic | Non-invasive LRTI Value (N=1593) | Invasive LRTI Value (N=1039) | P value |
|---|---|---|---|---|
| CURB 65 Category | 0-1 (Mild) % (n) | 38.7% (616) | 33.4% (347) | <0.001 † |
| | 2 (Moderate) % (n) | 31.2% (497) | 28.8% (299) | |
| | 3-5 (Severe) % (n) | 30.1% (480) | 37.8% (393) | |
| Admitted ICU | yes % (n) | 14.2% (226) | 10.2% (106) | 0.0026 † |
| Intubation | yes % (n) | 9.4% (150) | 7.2% (75) | 0.054 † |
| Received Ionotropes | yes % (n) | 11.0% (176) | 7.9% (82) | 0.0088 † |
| Death Within 30 Days | yes % (n) | 11.9% (189) | 15.4% (160) | 0.0097 † |

†, Fisher's exact test (categorical)
An adjusted P value of 0.01 may be considered significant.

In the LRTI cohort invasive disease is more severe of presentation, and has a higher crude death rate at 30 days. These trends are broadly in line with the comparison of invasive to non-invasive disease of all clinical presentations in table 2 of the main paper. In contrast to the main paper, invasive disease patients are less frequently admitted to ICU, intubated, or receive ionotropes however there is a potential survival bias in this finding.

*Supplementary table 4: Adjusted Cox proportional hazard models for the outcomes of death within 30 days and discharge from hospital within 30 days (censored for death) in the subgroup of patients with LRTI.*

| | | Death within 30 days (Adjusted) (N=348) | | Discharge within 30 days (Adjusted) (N=2204) | |
|---|---|---|---|---|---|
| Characteristic | Subgroup | HR [95% CI] | P value | HR [95% CI] | P value |
| Invasive Status | Non-invasive LRTI | ref | 0.044 | ref | 0.171 |
| | Invasive LRTI | 1.24 [1.01 – 1.54] | | 1.06 [0.97 – 1.16] | |
| Age Category | 18-34 | ref | <0.001 | ref | <0.001 |
| | 35-49 | 1.14 [0.48 – 2.72] | | 0.85 [0.72 – 0.99] | |
| | 50-64 | 2.87 [1.36 – 6.05] | | 0.68 [0.58 – 0.79] | |
| | 65-74 | 4.72 [2.27 – 9.80] | | 0.63 [0.54 – 0.74] | |
| | 75-84 | 6.40 [3.11 – 13.21] | | 0.52 [0.44 – 0.61] | |
| | 85+ | 11.74 [5.71 – 24.11] | | 0.41 [0.35 – 0.50] | |
| Gender | Female | ref | 0.001 | ref | 0.025 |
| | Male | 1.43 [1.15 – 1.76] | | 0.91 [0.83 – 0.99] | |
| Any Chronic Lung Disease | no | ref | 0.582 | ref | 0.102 |
| | yes | 1.06 [0.86 – 1.31] | | 1.08 [0.99 – 1.17] | |

P values calculated using Likelihood ratio test (II)

In the LRTI cohort invasive disease status is associated with an increased risk of death, similar to the findings of the main paper. With regards to discharge the benefit of non-invasive disease is reduced in the LRTI cohort compared to that in the main paper which includes all presentations. This suggests the effect of clinical presentation and discharge is interacting with the effect of invasive disease status on discharge.

### Survival analysis: Invasive pneumococcal disease stratified by clinical presentation

*Supplementary table 5: A set of unadjusted Cox proportional hazards models relating a range of predictors to the outcome of death within 30 days in the LRTI, Meningitis, and Other presentation subgroups of invasive disease. For clarity confidence intervals are not shown.*

| | | LRTI (Unadjusted) | | Meningitis (Unadjusted) | | Other (Unadjusted) | |
|---|---|---|---|---|---|---|---|
| Characteristic | Subgroup | HR | P value | HR | P value | HR | P value |
| Age | | 1.05 | <0.001 | 1.02 | 0.166 | 1.04 | <0.001 |
| Gender | Female | ref | 0.811 | ref | 0.043 | ref | 0.204 |
| | Male | 1.04 | | 2.54 | | 0.73 | |
| Smoker | Non-smoker | ref | 0.006 | ref | 0.413 | ref | 0.229 |
| | Ex-smoker | 1.85 | | 2.01 | | 1.59 | |
| | Current | 1.26 | | 1.27 | | 1.08 | |
| Alcohol Abuse | no | ref | 0.102 | ref | 0.821 | ref | 0.161 |
| | yes | 1.63 | | 0.80 | | 1.63 | |
| CCI Category | None (0) | ref | <0.001 | ref | 0.004 | ref | <0.001 |
| | Mild (1-2) | 1.91 | | 0.99 | | 3.64 | |
| | Moderate (3-4) | 9.71 | | 0.33 | | 12.72 | |
| | Severe (5+) | 15.55 | | 4.24 | | 14.52 | |
| Any Chronic Lung Disease | no | ref | 0.081 | ref | 0.306 | ref | 0.621 |
| | yes | 1.32 | | 1.67 | | 1.13 | |
| Any Chronic Heart Disease | no | ref | <0.001 | ref | 0.829 | ref | <0.001 |
| | yes | 3.70 | | 1.11 | | 2.47 | |
| COPD | no | ref | <0.001 | ref | 0.045 | ref | 0.100 |
| | yes | 1.86 | | 3.15 | | 1.55 | |
| PPV23 Vaccination | no | ref | <0.001 | ref | 0.740 | ref | 0.057 |
| | yes | 1.84 | | 0.83 | | 1.60 | |
| White Cell Count | | 0.97 | 0.004 | 1.00 | 0.782 | 0.98 | 0.137 |
| CRP | | 1.00 | 0.747 | 1.00 | 0.091 | 1.00 | 0.622 |

P values calculated using Likelihood ratio test (II)

These unadjusted hazard ratios demonstrate that the factors that influence survival in meningitis are fundamentally different to those that determine survival in the other categories of presentation. This evidence is suggestive of a strong effect of gender on survival in pneumococcal meningitis, whereas this is not the case for other invasive pneumococcal disease. In contrast, smoking status is related to survival in LRTI but not other presentations. Whilst chronic heart or lung disease, and PPV23 vaccination are associated with poorer survival in LRTI, these patterns are not present for the meningitis cohort. These associations are unadjusted and many of the factors in this table are conflated with age, so these hazard rates should not be over interpreted. This comparison serves mainly to demonstrate that the factors influencing survival are different in pneumococcal meningitis compared to other types of invasive pneumococcal disease, and to caution against over-interpretation of adjusted survival models which compare these groups.

*Supplementary figure 1: Kaplan-Meier survival curves for death and discharge within 30 days in the subgroup of patients with invasive pneumococcal disease stratified by the clinical presentation. Subfigures show numbers of patients at risk in each category. Black dots show points at which observations were censored.*

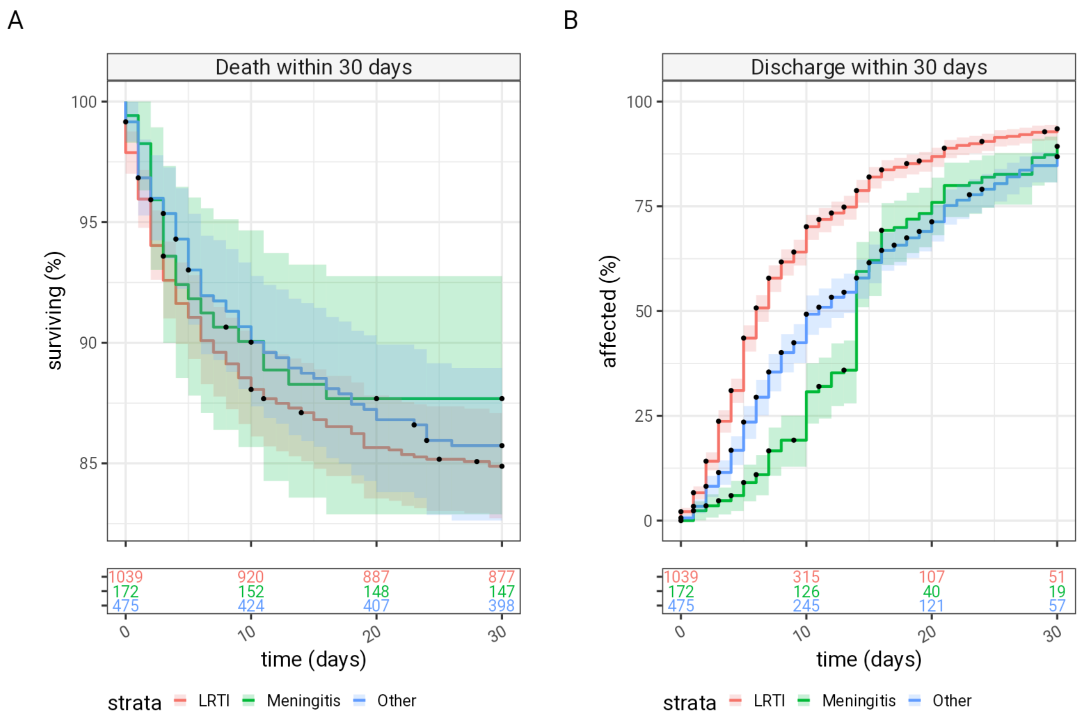


Despite the caution above, a survival analysis comparing clinical presentation is moderately informative. There is a tendency towards poorer 30 day survival with meningitis, however it is difficult to disentangle the effect of clinical presentation from that of gender. From this evidence, we could infer that men with pneumococcal meningitis perform significantly worse than other groups. The increase in time to discharge seen in meningitis and other non-LRTI presentations is striking in the adjusted models (supplementary table 2) but the Kaplan Meier curves (supplementary figure 1) suggest that this is not a proportional hazard and direct comparison in this manner may not be reliable, however the finding that meningitis cases stay in hospital longer that LRTI cases is unsurprising, and likely represents a real phenomenon.

*Supplementary table 6: Adjusted Cox proportional hazard models for the outcomes of death within 30 days and discharge from hospital within 30 days (censored for death) in the subgroup of patients with invasive pneumococcal disease.*

| | | Death within 30 days (Adjusted) (N=245) | | Discharge within 30 days (Adjusted) (N=1358) | |
|---|---|---|---|---|---|
| Characteristic | Subgroup | HR [95% CI] | P value | HR [95% CI] | P value |
| Clinical Syndrome | LRTI | ref | 0.194 | ref | <0.001 |
| | Meningitis | 1.43 [0.90 – 2.28] | | 0.44 [0.36 – 0.53] | |
| | Other | 0.88 [0.66 – 1.18] | | 0.58 [0.51 – 0.66] | |
| Gender | Female | ref | 0.219 | ref | 0.082 |
| | Male | 1.17 [0.91 – 1.51] | | 0.91 [0.82 – 1.01] | |
| Age Category | 18-34 | ref | <0.001 | ref | 0.043 |
| | 35-49 | 1.38 [0.48 – 3.98] | | 0.92 [0.74 – 1.14] | |
| | 50-64 | 1.21 [0.39 – 3.81] | | 0.95 [0.71 – 1.25] | |
| | 65-74 | 1.42 [0.45 – 4.48] | | 0.89 [0.65 – 1.20] | |
| | 75-84 | 1.67 [0.53 – 5.28] | | 0.77 [0.56 – 1.06] | |
| | 85+ | 3.14 [1.00 – 9.87] | | 0.67 [0.48 – 0.93] | |
| CCI Category | None (0) | ref | <0.001 | ref | <0.001 |
| | Mild (1-2) | 1.54 [0.57 – 4.10] | | 0.72 [0.57 – 0.92] | |
| | Moderate (3-4) | 4.33 [1.61 – 11.67] | | 0.70 [0.54 – 0.92] | |
| | Severe (5+) | 5.67 [2.12 – 15.15] | | 0.58 [0.44 – 0.76] | |

P values calculated using Likelihood ratio test (II)

An adjusted Cox proportional hazard model assessing risk of death within 30-days of admission comparing outcome following LRTI and meningitis shows an non-significant trend to worse outcome with meningitis, however this should be interpreted with caution as this one model encompasses two diverse patient groups with different associations.

### Significance testing: Serotype distributions in invasive pneumococcal disease

In the subgroup of invasive pneumococcal disease we have information on the causative serotype. This is presented in a descriptive form in the main paper. A more formal comparison testing against the null hypothesis that serotype distribution is independent of clinical presentation follows:

*Supplementary figure 2: Serotype distributions for the subgroup of patients with invasive pneumococcal disease, for whom the pneumococcal serotype was known, binary comparison between the 3 clinical presentation subgroups. Confidence intervals on the proportions are estimated at the 0.999 level to account for the implicit multiple hypothesis testing. Significant divergences of the frequency of individual serotypes in each clinical presentation would be seen as comparator proportions being different compared to reference proportions and the shaded confidence limits not intersecting the line y=x.*

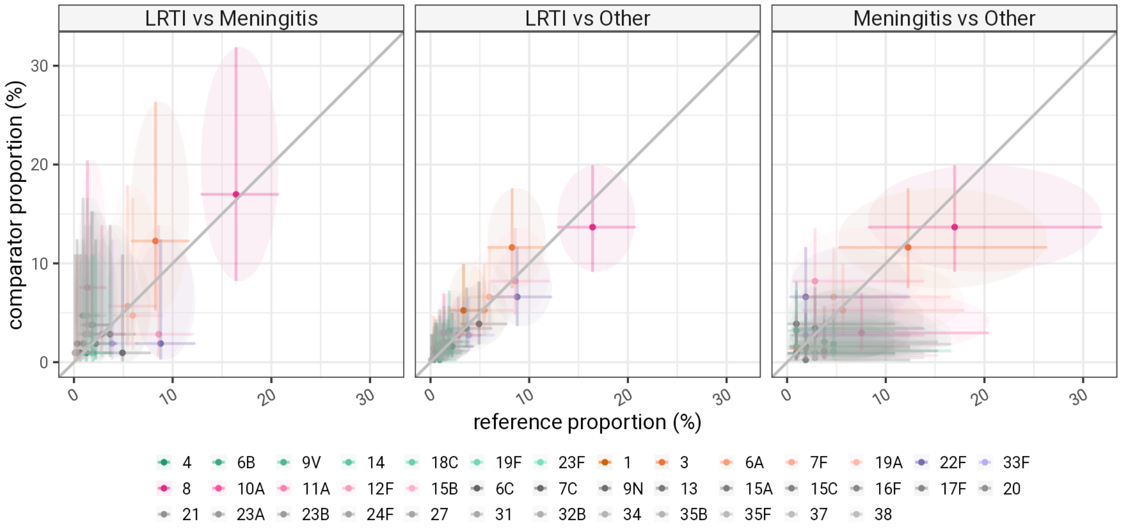


Neither direct comparison of the proportions of individual serotypes, for each clinical presentation (supplementary figure 2) or the observed versus expected counts of serotypes in each clinical presentation (supplementary figure 3) show clear significant differences between the serotype distribution in different clinical presentations. The vast majority of serotypes are observed in all groups within the expected 95% confidence limits with only the combination of serotype 10A and meningitis being present more than expected at 95%, but not at 99.9%. We are testing at least 50 multiple hypotheses simultaneously and hence observed values would only be interpreted as significant outliers if they were outside the 99.9% confidence limits. In this data all observations are consistent with the null hypothesis, at this level of significance (P<0.001).

*Supplementary figure 3: Observed versus expected serotype distributions for the IPD cohort. Expected serotype distributions were simulated assuming the null hypothesis that the serotype distribution is independent of clinical presentation, and using the marginal serotype and clinical presentation probabilities. 10000 bootstrap simulations of 1501 patients provide 95% (black bar) and 99.9% (grey bar) confidence intervals for the expected number of observations for each combination of serotype and clinical presentation. Observed frequencies of each serotype and clinical presentation are shown as red bars. Few observations fall outside of 95% confidence limits (Meningitis+10A, high; Meningitis+22F, low; Meningitis+1, low) and none outside 99.9% confidence limits, which adjust for implicit multiple hypothesis testing. In cases that are outside 95% confidence limits, effect sizes are small representing fewer than 10 cases over a 16 year period.*

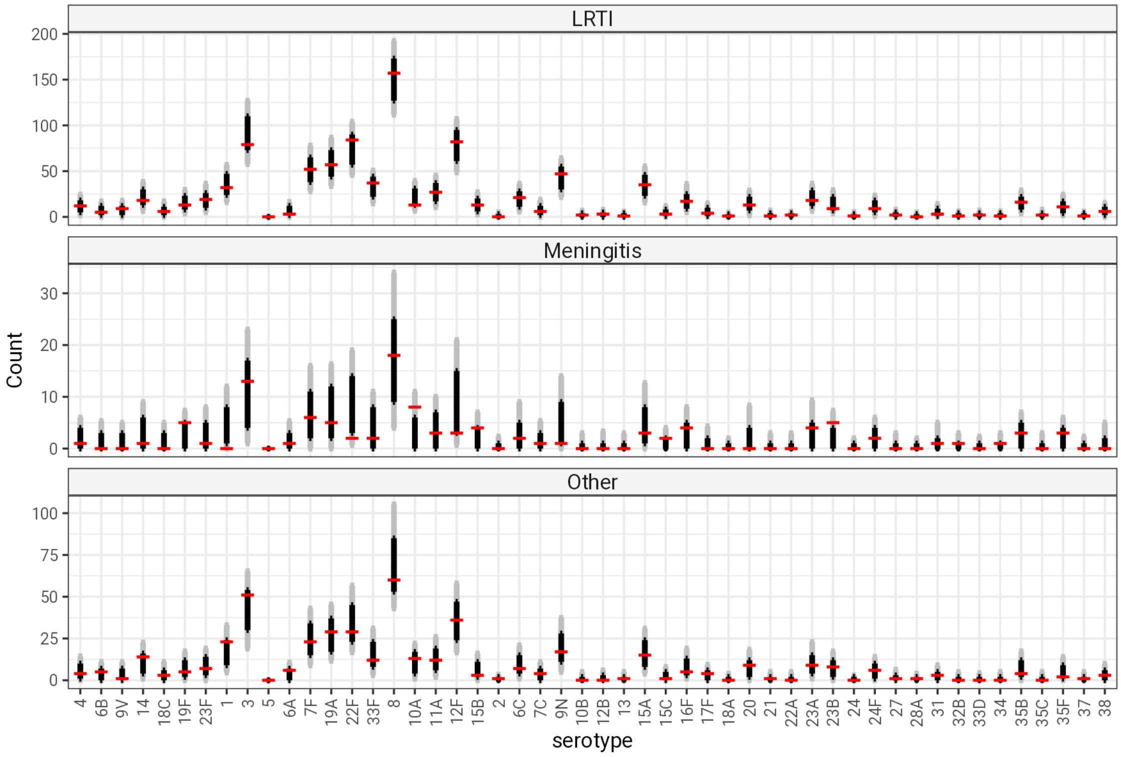
